# Are we there yet? An adaptive SIR model for continuous estimation of COVID-19 infection rate and reproduction number in the United States

**DOI:** 10.1101/2020.09.13.20193896

**Authors:** Mark B Shapiro, Fazle Karim, Guido Muscioni, Abel Saju Augustine

## Abstract

**Background:** The dynamics of the COVID-19 epidemic vary due to local population density and policy measures. When making decisions, policy makers consider an estimate of the effective reproduction number ℛ_t_ which is the expected number of secondary infections by a single infected individual.

**Objective:** We propose a simple method for estimating the time-varying infection rate and reproduction number ℛ_t_.

**Methods:** We use a sliding window approach applied to a Susceptible-Infectious-Removed model. The infection rate is estimated using the reported cases for a seven-day window to obtain continuous estimation of ℛ_t_. The proposed adaptive SIR (*aSIR*) model was applied to data at the state and county levels.

**Results:** The *aSIR* model showed an excellent fit for the number of reported COVID-19 positive cases, a one-day forecast MAPE was less than 2.6% across all states. However, a seven-day forecast MAPE reached 16.2% and strongly overestimated the number of cases when the reproduction number was high and changing fast. The maximal ℛ_t_ showed a wide range of 2.0 to 4.5 across all states, with the highest values for New York (4.4) and Michigan (4.5). We demonstrate that the *aSIR* model can quickly adapt to an increase in the number of tests and associated increase in the reported cases of infections. Our results also suggest that intensive testing may be one of the effective methods of reducing ℛ_t_.

**Conclusion:** The *aSIR* model provides a simple and accurate computational tool to obtain continuous estimation of the reproduction number and evaluate the efficacy of mitigation measures.

## Introduction

We are in the middle of a global COVID-19 pandemic caused by the SARS-CoV-2 virus. As of September 2, 2020, over 6 million individuals in the United States have been reported positive for SARS-CoV-2. Modeling studies are key for understanding factors that drive the spread of the disease and for developing mitigation strategies. Early modeling efforts forecasted very large numbers of infected individuals which would overwhelm healthcare systems in many countries [1–3]. These forecasts served as a call to action for policy makers to introduce policy measures including social distancing, travel restrictions, and eventually lockdowns to avoid the predicted catastrophe [4–6]. The mitigating policy measures have been successful in changing the dynamics of the epidemic and “flattening the curve” so that fewer people needed to seek treatment at any given time and as such not overwhelm the healthcare system.

One of the most fundamental metrics that describes the epidemic’s dynamics is the reproduction number ℛ_t_ which is the expected number of secondary infections by a single infectious individual [7]. The idea that the course of an epidemic is determined by the rate of contact between susceptible and infectious individuals was proposed by William Hamer in 1906 [8]. Later, Kermack and McKendrick [9] showed that epidemics stop not when there are no susceptible individuals left, but rather when each infected individual can infect on average fewer than one more person. The reproduction number ℛ_t_ depends on three factors: 1) the likelihood of infection per contact, 2) the period during which infectious individuals freely interact with those susceptible to contract the disease, and 3) rate of contact. The likelihood of infection per contact (factor 1) is determined by pathogen virulence and also by protective measures such as social distancing or wearing masks. Free interactions between infectious and susceptible individuals (factor 2) occur until the infectious individual is self-quarantined or hospitalized, either when the individual tests positive or symptoms become severe. Finally, the rate of contact (factor 3) is strongly affected by public health measures to mitigate risk [10], such as lockdowns during the COVID-19 epidemic. Thus, the reproduction number is determined by the biological properties of the pathogen and multiple aspects of social behavior. When ℛ_t_>1 the number of cases is expected to grow exponentially. The epidemic is contained when ℛ_t_ decreases and remains below 1. Real-time estimation of ℛ_t_ is critical for determining the effect of implemented mitigation measures and planning for the future.

We propose a method for the continuous estimation of infection rate and reproduction number ℛ_t_ that reflects the effects of mitigation measures as well as immunity acquired by those who recover from the disease. We estimate ℛ_t_ using a Susceptible-Infectious-Removed (SIR) model [9] that describes the dynamics of population compartments as follows: individuals start as Susceptible, are infected with the virus and become Infectious, and then move to the Removed compartment once they are quarantined or hospitalized, recover, or die. The SIR model is one of the simplest epidemiological models that still captures the main properties of an epidemic [11,12] and it has been widely used in epidemic modeling studies. In the majority of SIR modeling studies, the model parameters were constant. An SIR model with constant parameters, however, cannot be applied to the COVID-19 epidemic because various mitigating measures were introduced as the epidemic progressed. The effect of policy changes on COVID-19 dynamics has been modeled using a combination of an SIR model and Bayesian inference [13,14]. In these modeling studies, the infection spreading rate was assumed to be piece-wise linear between the three dates of policy changes. In another approach, continuous estimation of the reproduction number and the effect of mitigation measures were obtained based on estimates of the distribution of the serial interval between the symptom onset in the primary and secondary cases [15–17]. The Bayesian inference methods as well as methods based on estimations of the serial interval include multiple parameters whose values are not estimated from the data. In contrast, we propose an adaptive SIR model (*aSIR*) in which only one parameter, the removal rate, is taken from the literature, while the second parameter, the infection rate, is continuously estimated from the data using a sliding window. A continuous estimate of the reproduction number ℛ_t_ is then calculated using the infection rate estimate. The SIR model is described as a system of differential equations, and the key idea in our proposed method is that the initial conditions for each window are taken as values estimated for the previous window. The only additional hyperparameter is the length of the sliding window. The proposed method retains the conceptual and computational simplicity of SIR-type models and can be easily extended through the introduction of additional compartments supported by data.

### Data

The data on daily and cumulative confirmed cases between February 29, 2020 and September 2, 2020 were obtained from John Hopkins University (JHU) and the dates of interventions by states (e.g. state of emergency, stay-at-home order) were obtained from Wikipedia (Wikipedia, 2020). The JHU data were available at two levels of aggregation: county and state. JHU considers many sources for reporting these data; county level information is extracted from the website of the state’s departments of health (Johns Hopkins University, 2020) and state level data are extracted directly from the Centers for Disease Control website.

### Model

The SIR model is a system of ordinary differential equations:

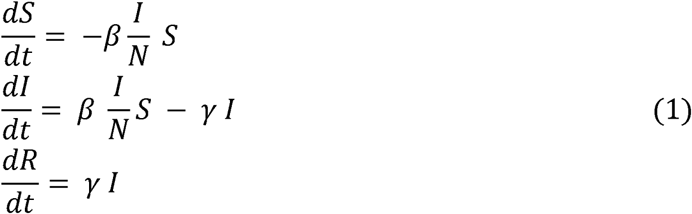

Here, *S* is the number of susceptible individuals; *I* is the number of infectious individuals who freely interact with others and can transmit the infection; *R* is the number of individuals removed from the other two compartments because they are quarantined or hospitalized, recover and acquire immunity, or die. Several COVID-19 government data sources provide the daily number of newly confirmed cases as well as a cumulative number of confirmed cases. Careful consideration is required to determine if these numbers should be attributed to the *I* or *R* compartment. In the US, once an individual has been confirmed COVID-19 positive that person is expected to be either self-isolated or hospitalized. Therefore, we assigned the data on confirmed cases to the *R* compartment, and we fit the model on the cumulative number of confirmed cases.

The infection rate *β* is *β = p* * *c*, where *p* is the probability of infection during contact with an infectious individual, and *c* is the average number of contacts per day. We have no data that would allow us to estimate *p* and *c* separately, so we directly estimate *β* as is usually done when using SIR models.

The removal rate *γ* determines the rate with which the infected are removed from *I* to the *R* compartment. In the context of the COVID-19 epidemic, *γ* is determined by the time it takes for severe symptoms to appear so the person gets tested and is self-quarantined or hospitalized. Therefore, we will assume the duration of the infectious period as the average time it takes for the infected person to become isolated, not the overall time to recover. We assume that the person is infectious from the day they get infected before the symptoms appear [18–20]. The average time to develop symptoms has been reported as 5 to 6 days [21–23]. We assume the infectious period before developing severe symptoms is 6 days, so *γ* = 1/6.

### Time-variant Parameter Estimation

The *aSIR* model contains two parameters, *β* and *γ*, with *γ =* 1/6 taken from the literature and *β* estimated using the reported data for each region of interest. The time-variant *β*(*t*) was estimated using a sliding window of *τ* =7 days and step of *s* =1 day, with the estimated values for *S* and *I* from the previous window used as the initial conditions for the next window. The reproduction number was calculated as ℛ_t_ (*t*) *= β*(*t*)*/γ*.

1. For the first window, we determined the date when the number of confirmed cases began to increase exponentially. This is important because for many states or counties, very few confirmed cases were initially reported for a number of days or even weeks, which suggests that either the epidemic had not started or the true number of infected people was not known. It is not reasonable to apply a SIR model for this initial period. We took the onset of the epidemic as the first of the four consecutive days in which the number of reported confirmed cases rose in at least three days. The initial conditions for system (1) for window 0 were *S*_*0*_*(0) = N*, where N is the population in the region of interest, *I*_*0*_*(0)* = 1, and *R*_*0*_*(0)* = 0. Infection rate *β*_*i*_ and *S(t), I(t)* for t ∈ [0, *τ*−1] were estimated given the initial conditions and actual *R*.
2. Slide the window by *s*=1 point. For the new *i+1* window, take the initial conditions as the estimated values from the previous window *S*_*i+1*_*(0) = S*_*i*_*(s), I*_*i+1*_*(0)= I*_*i*_*(s)*, and actual *R*_*i+1*_*(0)=R(s)*. Use actual values of *R*(*t*), estimate infection rate *β*_*i+1*_ and *S*_*i+1*_(*t*), *I*_*i+1*_(*t*).
3. For each window, calculate ℛ_*t*.*i*_ *= β*_*i*_ */γ*, assign the ℛ_*t*.*i*_ to the last time point of the window. To get a smooth estimate of ℛ_*t*_ we used a rolling average of 5 points.

## Results

We fit the model for each state and county in the United States. The model performance was evaluated by calculating the quality of fit as the root mean squared error between the actual and fitted *P* data for all windows concatenated (*wRMSE*). The fit was excellent with *wRMSE* < 6 across all states. We also calculated a 1-day forecast, 3-day forecast, and 7-day forecast of *R* after each window (Fig. 1A). The mean absolute prediction error (MAPE) for the forecasts is given in Table 1. The 1-day forecast error did not exceed 2.6% across all states while the 7-day forecast error was large and reached 16.2% for the state of New York. In particular, the 7-day forecast strongly overestimated the number of cases when the reproduction number was high and changing fast (Fig. 1).

**Table 1.** Reproduction numbers ℛ_*t*_ and forecast accuracy for 50 states.

| **State** | **$\mathcal{R}_t$**<br>**Max** | **MAPE**<br>**1-Day**<br>**Forecast** | **MAPE**<br>**3-Day**<br>**Forecast** | **MAPE**<br>**7-Day**<br>**Forecast** |
| --- | --- | --- | --- | --- |
| Alabama | 2.9 | 1.5% | 4.2% | 10.0% |
| Alaska | 2.8 | 1.6% | 3.7% | 11.3% |
| Arizona | 3.3 | 1.3% | 2.9% | 10.1% |
| Arkansas | 2.8 | 1.5% | 4.0% | 12.6% |
| California | 2.5 | 1.7% | 2.9% | 6.3% |
| Colorado | 2.6 | 1.1% | 2.9% | 7.3% |
| Connecticut | 4.1 | 2.0% | 3.1% | 9.3% |
| Delaware | 2.4 | 1.7% | 2.9% | 7.9% |
| District of Columbia | 2.1 | 0.8% | 1.8% | 4.4% |
| Florida | 3.6 | 2.0% | 4.4% | 9.3% |
| Georgia | 3.0 | 1.8% | 3.7% | 7.4% |
| Hawaii | 2.7 | 2.0% | 3.6% | 9.7% |
| Idaho | 3.4 | 2.4% | 4.8% | 13.6% |
| Illinois | 4.0 | 1.3% | 2.5% | 8.5% |
| Indiana | 3.8 | 1.4% | 4.0% | 10.4% |
| Iowa | 2.8 | 1.8% | 3.6% | 8.0% |
| Kansas | 3.0 | 1.6% | 3.5% | 8.6% |
| Kentucky | 3.0 | 2.6% | 4.9% | 11.2% |
| Louisiana | 3.7 | 1.8% | 4.0% | 12.1% |
| Maine | 2.0 | 1.2% | 2.8% | 6.7% |
| Maryland | 3.3 | 1.2% | 2.8% | 6.2% |
| Massachusetts | 3.4 | 1.3% | 3.6% | 9.7% |
| Michigan | 4.5 | 1.6% | 3.5% | 12.8% |
| Minnesota | 2.7 | 1.3% | 2.9% | 8.0% |
| Mississippi | 2.9 | 1.2% | 3.0% | 9.3% |
| Missouri | 3.6 | 1.8% | 3.3% | 11.4% |
| Montana | 3.3 | 1.6% | 3.7% | 11.9% |
| Nebraska | 2.5 | 2.0% | 4.1% | 9.7% |
| Nevada | 2.9 | 2.3% | 3.8% | 10.0% |
| New Hampshire | 2.3 | 1.7% | 3.3% | 8.5% |
| New Jersey | 4.1 | 1.5% | 2.3% | 7.8% |
| New Mexico | 2.3 | 2.2% | 3.4% | 7.3% |
| New York | 4.4 | 1.5% | 4.2% | 16.2% |
| North Carolina | 3.2 | 1.3% | 2.4% | 7.2% |
| North Dakota | 2.4 | 1.8% | 4.8% | 12.6% |
| Ohio | 3.3 | 1.2% | 3.4% | 9.8% |
| Oklahoma | 3.1 | 1.4% | 3.5% | 10.2% |
| Oregon | 2.5 | 1.3% | 2.8% | 6.6% |
| Pennsylvania | 3.2 | 1.7% | 2.9% | 6.3% |
| Rhode Island | 2.4 | 1.5% | 3.1% | 6.8% |
| South Carolina | 3.5 | 2.1% | 4.3% | 10.6% |
| South Dakota | 2.1 | 1.3% | 3.2% | 8.7% |
| Tennessee | 3.5 | 2.2% | 4.8% | 12.5% |
| Texas | 3.6 | 2.0% | 3.9% | 9.3% |
| Utah | 3.2 | 1.4% | 3.1% | 8.3% |
| Vermont | 2.9 | 0.8% | 2.4% | 7.7% |
| Virginia | 2.5 | 1.1% | 2.1% | 5.1% |
| Washington | 3.0 | 2.0% | 4.8% | 8.6% |
| West Virginia | 3.5 | 1.6% | 3.9% | 14.0% |
| Wisconsin | 3.6 | 1.5% | 3.2% | 10.0% |
| Wyoming | 2.9 | 1.9% | 4.7% | 14.2% |

**Figure 1.**
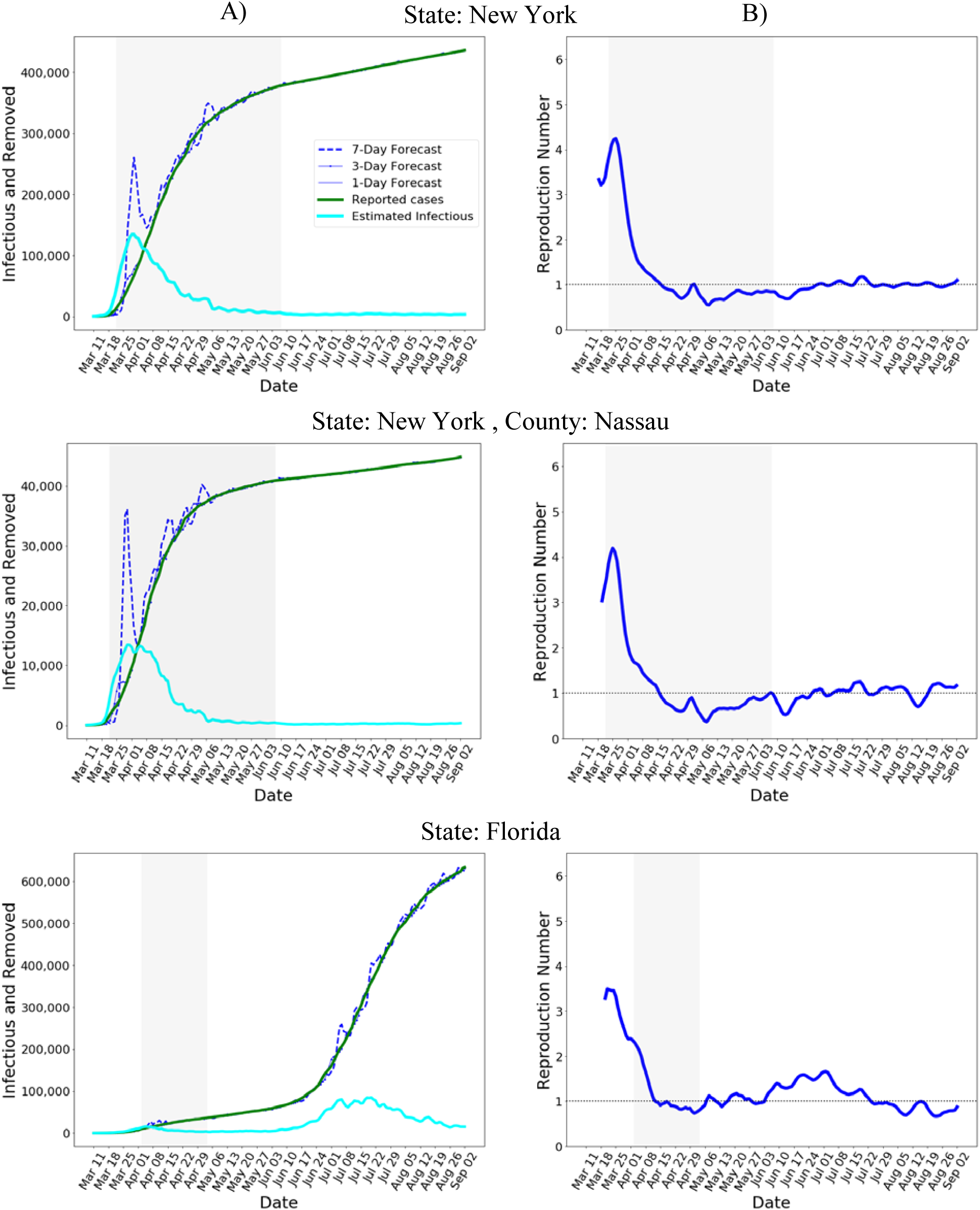
**A)** Estimated Infectious and forecasted Removed. **B)** Estimated reproduction number. The shaded region indicates the dates of the lockdown. While the 1-day and 3-day forecasts are accurate, the 7-day forecast exhibits large errors when >1 and is changing fast.

The estimated time course of ℛ_*t*_ for the state of New York and for Nassau county, one of the most affected in the beginning of COVID-19 epidemic, are shown in Figure 1. The estimated daily number of infectious individuals rapidly increased and then gradually declined after the lockdown was introduced on March 22, 2020 (Fig. 1A). The estimated reproduction number also declined after the lockdown began (Fig. 1B). The time course of ℛ_*t*_ shows weekly seasonality which likely reflects the effect of social interactions and possibly the effect of fluctuations in case reporting on weekdays vs. weekends. For New York state and Nassau county, ℛ_*t*_ exhibited an initial increase which may reflect the fact that the epidemic in the New York region was continuously seeded by travelers arriving to JFK airport until a ban on international travel was introduced on March 12, 2020. It may also reflect the fact that not all severe cases were initially recognized and reported as COVID-19. In Florida, ℛ_*t*_ decreased close to 1 by mid-April but then began increasing at the end of May (Fig. 1B). In June 2020, Florida authorities introduced more stringent measures to control the epidemic which is reflected in decreasing ℛ_*t*_ in the second half of July 2020. The opening of multiple states since June 2020 has been accompanied by ℛ_*t*_ rising above 1 (not shown here), and close monitoring of reproductive number is needed to contain another wave of the epidemic.

We also compared *aSIR* with the model by Cori and colleagues [15] implemented as R package *EpiEstim* and a model implemented by Kevin Systrom and Thomas Vladeck available from a popular and influential COVID-19 data tracking website *rt*.*live* (Fig. 2). In *EpiEstim*, we assumed equal probability of infection within the infectious period of 6 days, the estimate was smoothed with a 7-point rolling average window, same as in *aSIR*. While all three models show similar estimates when is close to 1, their estimates differ considerably in in the beginning of the epidemic. In particular, the *rt*.*live* model returned lower max than the other two models, and estimated that already decreased to 1 by the time the lockdown was announced in NY state on March 22, 2020 (Fig. 2, shaded region). The *EpiEstim* and the *aSIR* models estimated similar peak values of, and both models estimated that dereased close to 1 in the first week of April, 2020. Although both models show a rapidly decreasing in March, the *aSIR* model shows a lagged change. We are not aware of ground truth data however to determine which model produces a more accurate estimate.

**Figure 2.**
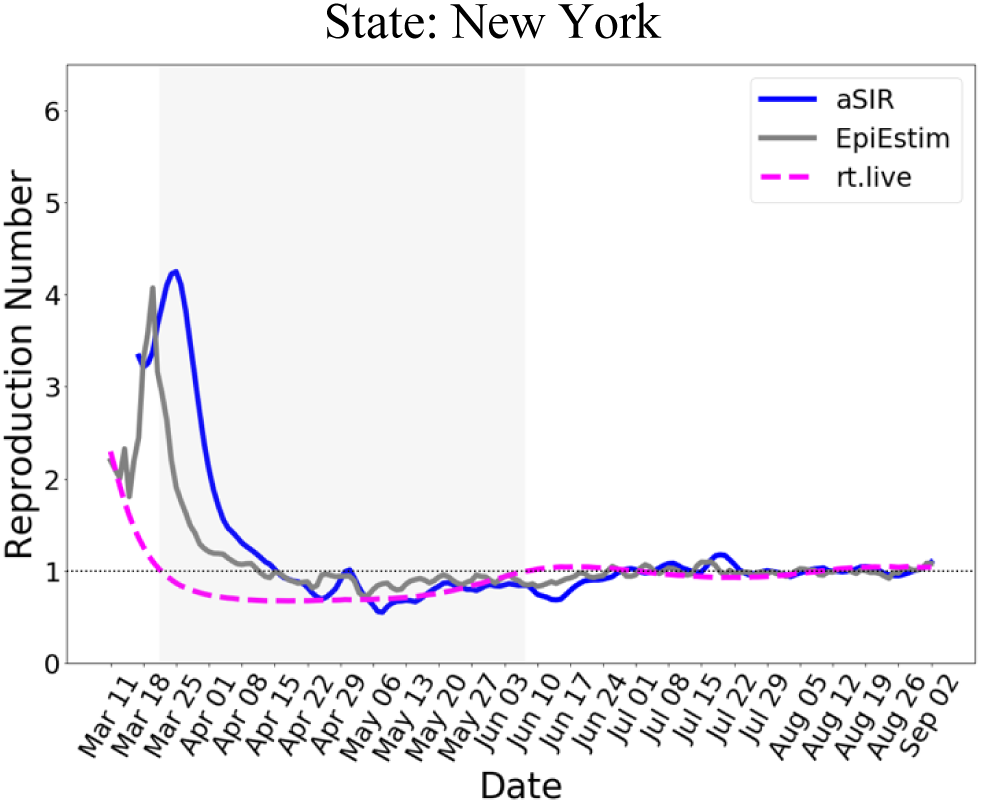
Comparison of models that generate continuous estimates. The three estimates differ widely in the beginning of the epidemic. In particular, the estimated by *rt*.*live* model decreased to 1 by the lockdown onset on March 22 (shaded region).

Next, we investigated the effect of an abrupt increase in testing on the estimate (Fig. 3). We assumed a step-wise 50% increase in testing that persisted after April 12 (Fig. 3, left panel). Both *aSIR* and *EpiEstim* models exhibited a spike in. It can also be argued that if testing increases then infectious individuals may be detected and quarantined sooner, resulting in a shorter infectious period and larger removal rate, in turn lowering. We did not model a possible increase in. Instead, we assumed that the underlying dynamics of the epidemic did not change, and within 2 weeks both models returned to the time course estimated without the testing increase.

**Figure 3.**
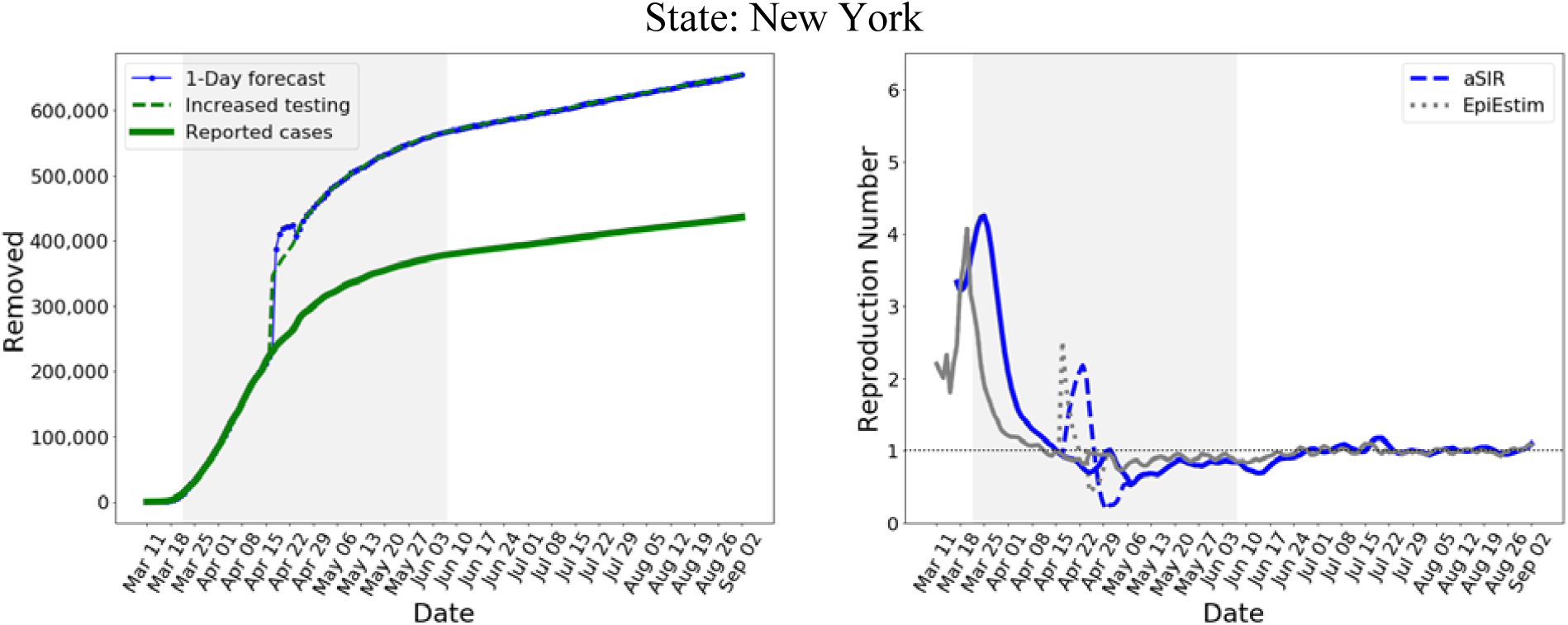
The effect of a step-wise 50% increase in testing (left panel, dashed line). The 1-day forecast by the *aSIR* model adapts within a week. For estimate, both *EpiEstim* and our *aSIR* models produced a spike followed by a decrease (right panel, dashed lines) before returning to the unperturbed time course (solid lines).

## Discussion

We have developed a simple approach to adaptively estimate time-varying parameters of the SIR model using reported data on the number of confirmed cases. This approach adds to the already large literature on COVID-19 modeling in two ways. First, we estimate the parameters of the SIR model using a sliding window of a limited duration, 7 days, to account for fast changes in transmissibility and contact patterns in response to changes in social behavior and government mitigation measures. The window duration is a hyperparameter that can be changed as needed, the trade-off being the accuracy of the parameter estimates versus rapid reaction to changes in the underlying epidemic. Because the proposed model is so simple, a number of scenarios can be explored as needed.

Second, we attribute the data on reported cases to the Removed compartment rather than Infectious. This modeling decision is based on the realities of the COVID-19 epidemic in the US where confirmed positive individuals are supposed to self-isolate or are hospitalized. Although these individuals remain infectious and can infect other family members or caretakers even when self-isolated or hospitalized, they are not freely interacting with the susceptible population as would be required to attribute them to the *I* compartment. It has been proposed to add a new X compartment in the SIR model to model symptomatic quarantined infectious individuals [24]. We have no data to independently estimate this additional parameter of quarantine rate, however. For the same reason, we did not use the Susceptible-Exposed-Infected-Removed (SEIR) model because we are not aware of reliable data about the duration of the exposed period during which an infected person is not yet infectious. Moreover, it has been found that the SIR model performed better than an SEIR model in representing the information contained in the confirmed-case data on COVID-19 [25].

The reported numbers of positive cases represent a fraction of infected individuals because of the limited availability of testing in March and April of 2020 with the result that only those who developed severe symptoms were tested. Up to 80% of the infected individuals may be asymptomatic or develop mild symptoms [26] and were not tested, so for that period our model applies only to the small sub-population who developed severe symptoms. This sub-population, however, is of particular interest because it represents those most at risk, and the reproduction number estimated from this limited data can be used to guide policy decisions aimed at protecting the most vulnerable population [27]. At the same time, as the numbers of the tested individuals increase, the short sliding window approach makes our model adaptable to an ever-larger proportion of the population (Fig. 3).

Across all US states, the maximal ℛ_*t*_ values of were estimated for New York (4.4) and Michigan (4.5) (Table 1) which is close to the mean value of 4.34 estimated for Italy [28] but higher than that obtained by a stochastic transmission model [29,30]. The wide range of maximal values of ℛ_*t*_ from 2.0 to 4.5 (Table 1) likely reflects the differences in contact rates due to population density [31,32]. Increased social distancing is required to contain the spread of the epidemic [33], with more stringent mitigation measures, including lockdown, considered necessary to decrease the contact rate in high-density states and counties. Another measure to lower ℛ_*t*_ is to increase the removal rate *γ* by intensive testing and quarantine of individuals tested positive. This targeted intervention would strongly decrease the interaction between the infectious and susceptible individuals and keep ℛ_*t*_ <1 until a vaccine becomes available. Our model will allow researchers as well as policy makers to monitor the reproduction number in different geographical regions of the US, better understand the effect of government policies on the dynamics of the epidemic, and develop further mitigation strategies as we continue to battle COVID-19 [34,35].

## Data Availability

The data on daily and cumulative confirmed cases between February 29, 2020 and September 2, 2020 were obtained from John Hopkins University. The dates of interventions by states were obtained from Wikipedia. State level data were extracted directly from the Centers for Disease Control website.
The code is available on github https://github.com/fazlekarim-anthem/aSIR

